# Predictors of Developing Renal Dysfunction Following Diagnosis of Transthyretin Cardiac Amyloidosis

**DOI:** 10.1101/2024.01.12.24301255

**Authors:** Malcolm L McDonald, Yosef Manla, Alice Sonnino, Mileydis Alonso, Radhika K Neicheril, Alejandro Sanchez, Gabrielle Lafave, Yelenis Seijo De Armas, Antonio Lewis Camargo, Dipan Uppal, Armaan Handa, David Wolinsky, Nina Thakkar Rivera, Mauricio Velez, David Baran, Jerry D. Estep, David Snipelisky

## Abstract

**Background:** In patients with transthyretin cardiac amyloidosis (ATTR-CA), renal dysfunction is a poor prognostic indicator. Limited data are available on variables that portend worsening renal function (wRF) among ATTR-CA patients.

**Objectives:** This study assesses which characteristics place patients at higher risk for the development of wRF (defined as a drop of ≥ 10% in GFR) within the first year following diagnosis of ATTR-CA.

**Methods:** We included patients with ATTR-CA (n=134) evaluated between 2/2016 and 12/2022 and followed for up to one-year at our amyloid clinic. Patients were stratified into two groups: a group with maintained renal function (mRF) and a group with wRF and compared using appropriate testing. Significant variables in the univariate analysis were included in the multivariable logistic regression model to determine characteristics associated with wRF.

**Results:** Within a follow-up period of 326±118 days, the median GFR% change measured -6% [-18%, +8]. About 41.8% (n=56) had wRF, while the remainder had mRF. In addition, in patients with no prior history of CKD, 25.5% developed de-novo CKD. On multivariable logistic regression, only NYHA class ≥III (OR: 3.9, 95% CI [1.6-9.3]), history of IHD (OR:0.3, 95% CI [0.1-0.7]), and receiving SGLT-2i (OR: 0.1, 95% CI [0.02-0.5]) were significant predictors of wRF.

**Conclusion:** Our study demonstrated that the development of new or worsening renal dysfunction is common following the diagnosis of ATTR-CA. Additionally, we identified worse NYHA class and no prior history of IHD as significant predictors associated with developing wRF, while receiving SGLT2i therapy appeared to be protective in this population.

## Introduction

Transthyretin cardiac amyloidosis (ATTR-CA) is a rare, progressive, and ultimately fatal disease that results from the misfolding and aggregation of amyloidogenic transthyretin (TTR) proteins in the myocardial extracellular space (1,2). ATTR-CA can result from an idiopathic subtype, also known as wild-type (wtATTR-CA), or a hereditary subtype (hATTR-CA) resulting from a pathogenic genetic mutation (1–3). Irrespective of the subtype, deposition of protein in patients with ATTR-CA not uncommonly extends beyond myocardial tissue to other organ systems. The kidney, nervous system, and gastrointestinal tract, among others, can suffer from protein deposition, leading to a wide array of phenotypic manifestations of the disease with an equally broad array of morbidity and disability burdens (1,3).

Direct renal involvement is primarily described in hATTR-CA, occurring to a much lesser extent in wtATTR-CA(1,3–5). Tissue deposition of misfolded ATTR in the glomeruli, renal vasculature, and interstitium is well-documented as a potential mechanism for worsening renal function (wRF) (3–5). However, the renal impacts of this systemic disease extend beyond local tissue effects. Through several complex physiological cascades, including neurohormonal activation, vascular non-compliance, poor tissue perfusion, and worsening autonomic dysfunction, the interplay between the failing heart and the kidney often results in further disease progression and end-organ decline (1).

Although renal dysfunction is a poor prognostic indicator among patients with ATTR-CA, most studies focus only on the hATTR-CA subset and assess characteristics associated with already established renal disease (4,6). Considering the morbidity and mortality risks associated with renal disease in this population, the aim of this study is to assess which clinical characteristics place patients at risk for development of wRF or progression of known disease following the diagnosis of ATTR-CA.

## Methods

### Study Population and Design

Consecutive patients evaluated in our amyloid clinic with a confirmed diagnosis of ATTR-CA between 2/2016 and 11/2022 were assessed. Patients included were diagnosed with either wtATTR or hATTR. Patients with light chain (AL) amyloidosis were excluded. Data on patient demographics, comorbidities, medical history, imaging and laboratory findings, and clinical presentation were collected retrospectively by review of the electronic medical record. Patients with incomplete data were excluded. This study was approved by the Cleveland Clinic Foundation Institutional Review Board.

### Study Variables and Definitions

ATTR-CA diagnosis was confirmed through verification of endomyocardial biopsy tissue samples or ^99^Tc-pyrophosphate scanning combined with single photon emission computed tomography (SPECT) imaging. The presence of AL amyloidosis was ruled out by laboratory testing and, if indicated, either bone marrow or endomyocardial biopsy. History of CKD was defined as having an estimated glomerular filtration rate of less than 60 ml/min/1.73m^2^ based on the Chronic Kidney Disease Epidemiology Collaboration (CKD-EPI) equation. Glomerular filtration rate (GFR) was recorded at the time of diagnosis and the last follow-up encounter (closest to one year). Given the considerable instability of the GFR value on measurement and variations in follow-up periods in studies evaluating renal outcomes, there is no concrete definition of rapid decline in renal function (4,6,7),. We defined wRF as a decline of ≥ 10% in GFR over time, a cut-off that has been previously defined and validated to predict cardiovascular and renal endpoints in other clinical studies (8). Accordingly, patients were stratified into two groups: patients with maintained renal function (mRF), which included patients with either an increase or decrease in GFR of up to 10% over the study period, and a group with wRF, defined as a drop of ≥ 10% in GFR over time. A new incidence of CKD was defined as having a measurement of eGFr of <60 ml/min/1.73m^2^ within the study period despite the patient having an eGFR ≥ 60 ml/min/1.73m^2^ at baseline.

### Statistical Analysis

Categorical variables were presented as frequencies and percentages and compared using a chi-square test. Normally distributed continuous variables were presented as mean and standard deviation and compared using t-tests or paired t-test, as appropriate. Non-normally distributed continuous variables were presented as medians and quartiles and compared using Wilcoxon rank-sum tests. The wRF and mRF groups were compared, and statistically significant (P<0.05) variables in the univariate analysis were included in the multivariable logistic regression model to determine characteristics associated with wRF. A significance level of 0.05 was assumed for all tests. Analysis was performed using JMP® Data Analysis (Software Version 16, SAS Institute Inc., Cary, NC, USA).

## Results

A total of 134 patients with a confirmed diagnosis of ATTR-CA were included in this study. Within a follow-up period of 326±118 days, there was an overall decrease in GFR at follow-up compared to baseline (58.4 vs. 53.7, P<0.001), and the median GFR% change over time measured -6% [-18%, +8]. When stratifying patients according to GFR change over time, 58.2% (n=78) had mRF, while the rest had wRF (41.8%, n=56) with a change over time in absolute GFR of -12.5 [-22.5, -8] and a median GFR% change over time of -22% [-33%, -15%]).

When comparing the mRF and the wRF groups, there were no differences in age (78 ± 7.7 vs. 78.8 ± 8.7, P=0.6) or rates of male gender (92.3% vs. 89.3%, P=0.5), however patients with wRF were more likely African American (AA) (26.8% vs. 10.3%, OR:3.2, 95% CI [1.2-8.2] P=0.01). Additionally, no difference in history of hypertension (P=0.9), diabetes mellitus (P=0.6), atrial fibrillation (P=0.7), or peripheral vascular disease (P=0.3) were noted. Patients in the wRF group were less likely to have a history of ischemic heart disease (IHD) (48.2% vs.71.8%, OR: 0.4 95% CI [0.2-0.8], P=0.01) or prior diagnosis of chronic kidney disease (CKD) (55.4% vs. 71.8%, OR:0.5. 95%CI [0.2-1] P=0.04). At the time of ATTR-CA diagnosis, there were no group differences in ejection fraction (P=0.8) or NT-proBNP levels (P=0.2) **(Table 1)**. Nonetheless, patients in the wRF group tended to present with NYHA class ≥III (66.1% vs. 47.4%, OR:2.1, 95% CI [1.1-4.4], P=0.03).

**Table 1.**
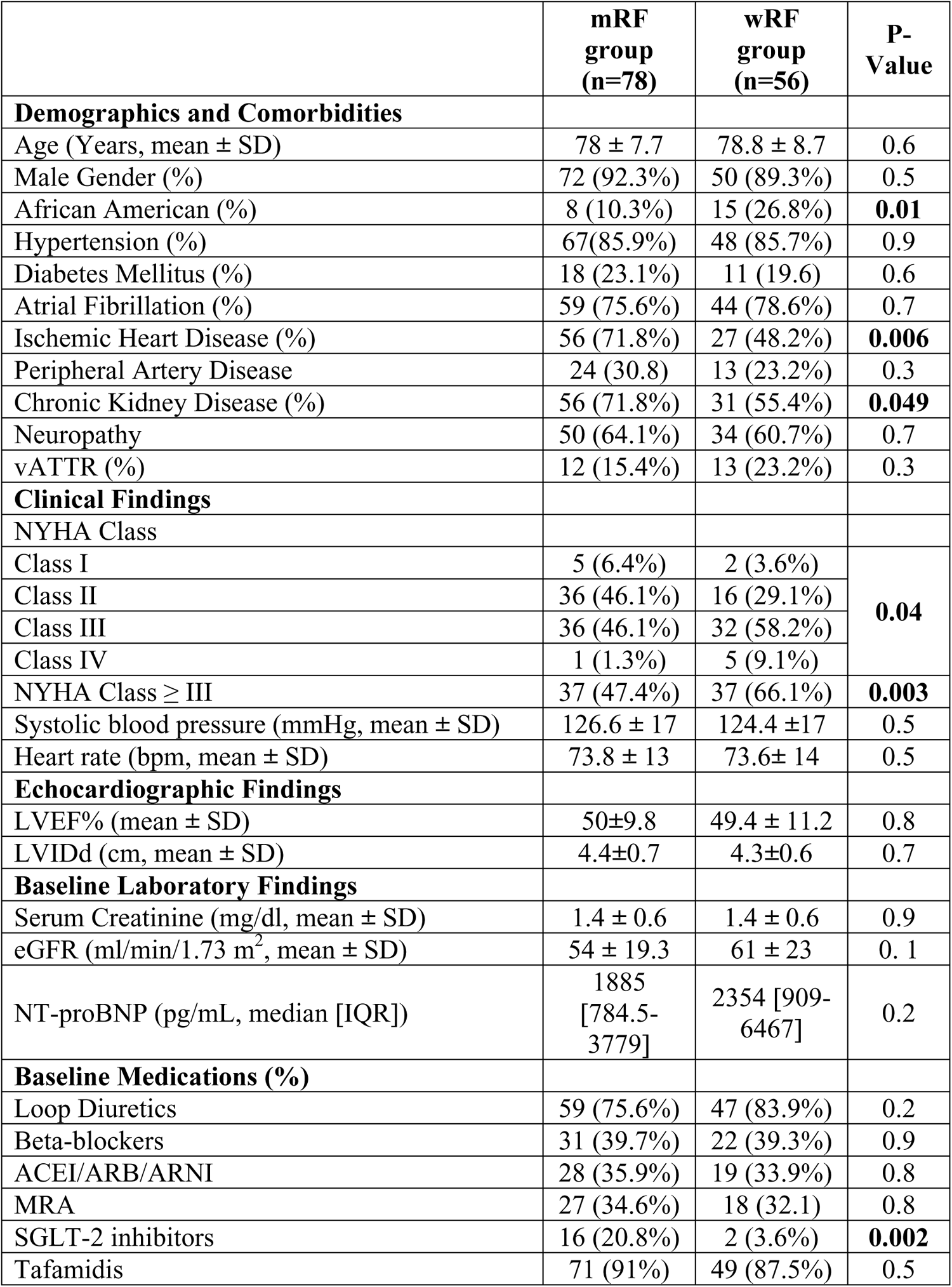
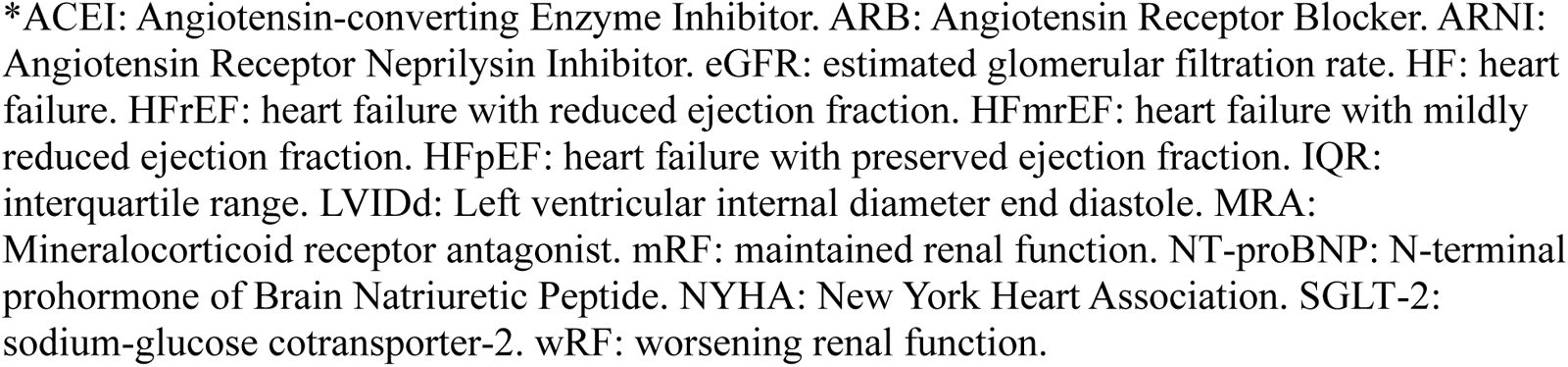
A Comparison of Baseline Clinical Characteristics of Patients Diagnosed with ATTR Cardiac Amyloidosis According to Their Renal Outcomes within One-year Follow-up.

|  | **mRF**<br>**group**<br>**(n=78)** | **wRF**<br>**group**<br>**(n=56)** | **P-**<br>**Value** |
| --- | --- | --- | --- |
| **Demographics and Comorbidities** |  |  |  |
| Age (Years, mean ± SD) | 78 ± 7.7 | 78.8 ± 8.7 | 0.6 |
| Male Gender (%) | 72 (92.3%) | 50 (89.3%) | 0.5 |
| African American (%) | 8 (10.3%) | 15 (26.8%) | **0.01** |
| Hypertension (%) | 67(85.9%) | 48 (85.7%) | 0.9 |
| Diabetes Mellitus (%) | 18 (23.1%) | 11 (19.6) | 0.6 |
| Atrial Fibrillation (%) | 59 (75.6%) | 44 (78.6%) | 0.7 |
| Ischemic Heart Disease (%) | 56 (71.8%) | 27 (48.2%) | **0.006** |
| Peripheral Artery Disease | 24 (30.8) | 13 (23.2%) | 0.3 |
| Chronic Kidney Disease (%) | 56 (71.8%) | 31 (55.4%) | **0.049** |
| Neuropathy | 50 (64.1%) | 34 (60.7%) | 0.7 |
| vATTR (%) | 12 (15.4%) | 13 (23.2%) | 0.3 |
| **Clinical Findings** |  |  |  |
| NYHA Class |  |  |  |
| Class I | 5 (6.4%) | 2 (3.6%) | **0.04** |
| Class II | 36 (46.1%) | 16 (29.1%) |  |
| Class III | 36 (46.1%) | 32 (58.2%) |  |
| Class IV | 1 (1.3%) | 5 (9.1%) |  |
| NYHA Class ≥ III | 37 (47.4%) | 37 (66.1%) | **0.003** |
| Systolic blood pressure (mmHg, mean ± SD) | 126.6 ± 17 | 124.4 ± 17 | 0.5 |
| Heart rate (bpm, mean ± SD) | 73.8 ± 13 | 73.6 ± 14 | 0.5 |
| **Echocardiographic Findings** |  |  |  |
| LVEF% (mean ± SD) | 50±9.8 | 49.4 ± 11.2 | 0.8 |
| LVIDd (cm, mean ± SD) | 4.4±0.7 | 4.3±0.6 | 0.7 |
| **Baseline Laboratory Findings** |  |  |  |
| Serum Creatinine (mg/dl, mean ± SD) | 1.4 ± 0.6 | 1.4 ± 0.6 | 0.9 |
| eGFR (ml/min/1.73 m<sup>2</sup>, mean ± SD) | 54 ± 19.3 | 61 ± 23 | 0.1 |
| NT-proBNP (pg/mL, median [IQR]) | 1885<br>[784.5-3779] | 2354 [909-6467] | 0.2 |
| **Baseline Medications (%)** |  |  |  |
| Loop Diuretics | 59 (75.6%) | 47 (83.9%) | 0.2 |
| Beta-blockers | 31 (39.7%) | 22 (39.3%) | 0.9 |
| ACEI/ARB/ARNI | 28 (35.9%) | 19 (33.9%) | 0.8 |
| MRA | 27 (34.6%) | 18 (32.1) | 0.8 |
| SGLT-2 inhibitors | 16 (20.8%) | 2 (3.6%) | **0.002** |
| Tafamidis | 71 (91%) | 49 (87.5%) | 0.5 |

\*ACEI: Angiotensin-converting Enzyme Inhibitor. ARB: Angiotensin Receptor Blocker. ARNI: Angiotensin Receptor Neprilysin Inhibitor. eGFR: estimated glomerular filtration rate. HF: heart failure. HFrEF: heart failure with reduced ejection fraction. HFmrEF: heart failure with mildly reduced ejection fraction. HFpEF: heart failure with preserved ejection fraction. IQR: interquartile range. LVIDd: Left ventricular internal diameter end diastole. MRA: Mineralocorticoid receptor antagonist. mRF: maintained renal function. NT-proBNP: N-terminal prohormone of Brain Natriuretic Peptide. NYHA: New York Heart Association. SGLT-2: sodium-glucose cotransporter-2. wRF: worsening renal function.

Use of tafamidis at the time of diagnosis was high in both groups (91% vs. 87.5%, P=0.5), and patients who developed wRF were less likely to be on SGLT-2i (3.6% vs. 20.8%, OR:0.14, 95% CI [0.03-0.65], P=0.002). No differences were noticed in rates of prescription of other guideline-directed medical therapies between groups. Further, in a sub-analysis of patients with mRF, there was a trend towards a 45% slower decline in GFR% over time (median decline: 3.9% vs. 7.1%, P=0.08) among SGLT-2i receivers as compared to non-receivers. Among this subset of patients, no other statistical differences were identified between any of the remaining characteristics analyzed, including age, race, comorbidities, functional class, ATTR-CA type, and concomitant use of other pharmacologic agents.

On a multivariable logistic regression including AA race, history of CKD, history of IHD, NYHA class ≥III at presentation, and receiving SGLT-2i, only NYHA class ≥III (OR: 3.9, 95% CI[1.6-9.3]), history of IHD (OR: 0.3, 95% CI [0.1-0.7]), and SGLT-2i prescription (OR: 0.1, 95% CI [0.02-0.5]) were significant predictors of wRF (**Figure 1**).

**Figure 1.**
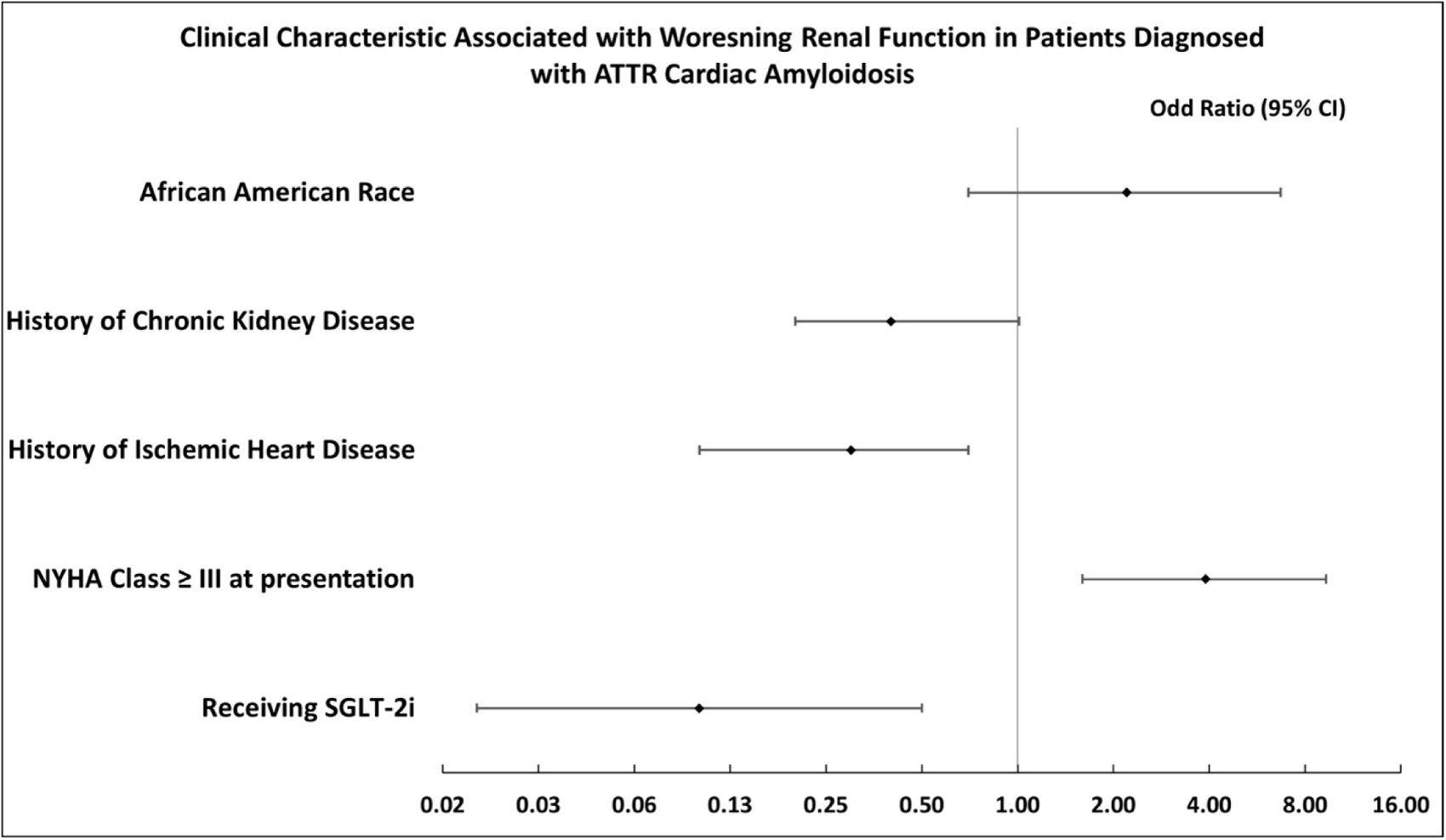
Clinical characteristics associated with worsening renal function in patients diagnosed with ATTR cardiac amyloidosis. *AA: African American, ATTR-CA: transthyretin cardiac amyloidosis, CKD: Chronic Kidney Disease, IHD: ischemic Heart Disease, NT-proBNP: N-terminal prohormone of Brain Natriuretic Peptide, NYHA: New York Heart Association, SGLT-2i: sodium-glucose cotransporter-2 inhibitors. wRF: worsening renal function.

In a sub-analysis of patients with no prior history of CKD (n=47), 25.5% (n=12) developed de novo CKD over the study period. Compared to those who maintained an eGFR≥ 60 during the study period, a higher proportion of patients with NYHA class ≥ III (75% vs. 40%, P=0.04), higher levels of NT-proBNP (3495 [1905-11431] vs. 972 [542.2-2512], P=0.001), higher serum Cr levels (1.1 ± 0.15 vs. 0.9 ± 0.14, P=0.001), lower LVEF (45±13 vs. 52±8.3, P=0.04), and higher rates of diuretic use (91.7% vs. 57.1%, P=0.03) were noted among those who developed CKD (**Table 2**).

**Table 2.**
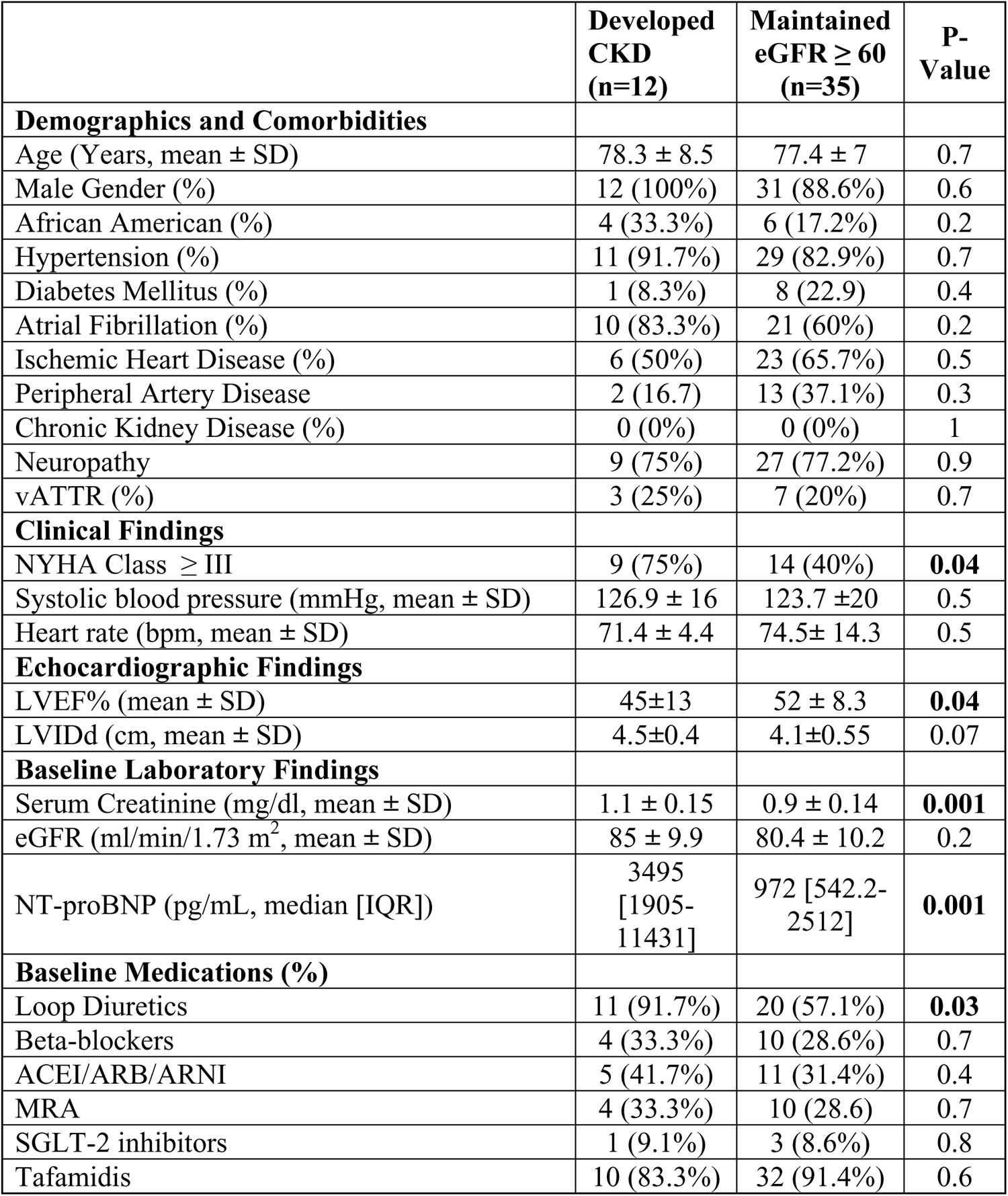
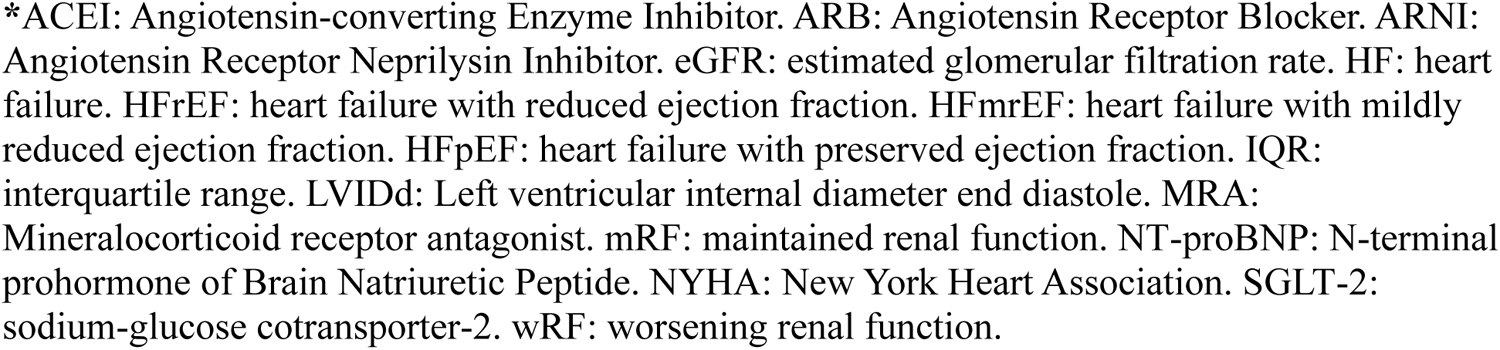
A Comparison of Baseline Clinical Characteristics of Patients Who Developed CKD During the vs. Those Who Maintained an eGFR ≥ 60 ml/min/1.7 m^2^ Within One-year Follow-up.

|  | **Developed**<br>**CKD**<br>**(n=12)** | **Maintained**<br>**eGFR ≥ 60**<br>**(n=35)** | **P-**<br>**Value** |
| --- | --- | --- | --- |
| **Demographics and Comorbidities** |  |  |  |
| Age (Years, mean ± SD) | 78.3 ± 8.5 | 77.4 ± 7 | 0.7 |
| Male Gender (%) | 12 (100%) | 31 (88.6%) | 0.6 |
| African American (%) | 4 (33.3%) | 6 (17.2%) | 0.2 |
| Hypertension (%) | 11 (91.7%) | 29 (82.9%) | 0.7 |
| Diabetes Mellitus (%) | 1 (8.3%) | 8 (22.9) | 0.4 |
| Atrial Fibrillation (%) | 10 (83.3%) | 21 (60%) | 0.2 |
| Ischemic Heart Disease (%) | 6 (50%) | 23 (65.7%) | 0.5 |
| Peripheral Artery Disease | 2 (16.7) | 13 (37.1%) | 0.3 |
| Chronic Kidney Disease (%) | 0 (0%) | 0 (0%) | 1 |
| Neuropathy | 9 (75%) | 27 (77.2%) | 0.9 |
| vATTR (%) | 3 (25%) | 7 (20%) | 0.7 |
| **Clinical Findings** |  |  |  |
| NYHA Class ≥ III | 9 (75%) | 14 (40%) | **0.04** |
| Systolic blood pressure (mmHg, mean ± SD) | 126.9 ± 16 | 123.7 ± 20 | 0.5 |
| Heart rate (bpm, mean ± SD) | 71.4 ± 4.4 | 74.5 ± 14.3 | 0.5 |
| **Echocardiographic Findings** |  |  |  |
| LVEF% (mean ± SD) | 45±13 | 52 ± 8.3 | **0.04** |
| LVIDd (cm, mean ± SD) | 4.5±0.4 | 4.1±0.55 | 0.07 |
| **Baseline Laboratory Findings** |  |  |  |
| Serum Creatinine (mg/dl, mean ± SD) | 1.1 ± 0.15 | 0.9 ± 0.14 | **0.001** |
| eGFR (ml/min/1.73 m<sup>2</sup>, mean ± SD) | 85 ± 9.9 | 80.4 ± 10.2 | 0.2 |
| NT-proBNP (pg/mL, median [IQR]) | 3495<br>[1905-11431] | 972 [542.2-2512] | **0.001** |
| **Baseline Medications (%)** |  |  |  |
| Loop Diuretics | 11 (91.7%) | 20 (57.1%) | **0.03** |
| Beta-blockers | 4 (33.3%) | 10 (28.6%) | 0.7 |
| ACEI/ARB/ARNI | 5 (41.7%) | 11 (31.4%) | 0.4 |
| MRA | 4 (33.3%) | 10 (28.6) | 0.7 |
| SGLT-2 inhibitors | 1 (9.1%) | 3 (8.6%) | 0.8 |
| Tafamidis | 10 (83.3%) | 32 (91.4%) | 0.6 |

\*ACEI: Angiotensin-converting Enzyme Inhibitor. ARB: Angiotensin Receptor Blocker. ARNI: Angiotensin Receptor Neprilysin Inhibitor. eGFR: estimated glomerular filtration rate. HF: heart failure. HFrEF: heart failure with reduced ejection fraction. HFmrEF: heart failure with mildly reduced ejection fraction. HFpEF: heart failure with preserved ejection fraction. IQR: interquartile range. LVIDd: Left ventricular internal diameter end diastole. MRA: Mineralocorticoid receptor antagonist. mRF: maintained renal function. NT-proBNP: N-terminal prohormone of Brain Natriuretic Peptide. NYHA: New York Heart Association. SGLT-2: sodium-glucose cotransporter-2. wRF: worsening renal function.

## Discussion

Our study found that worsening renal function following the diagnosis of ATTR-CA was quite common, with over 40% of patients developing new-onset or worsening of known renal disease within the first year following diagnosis of ATTR-CA and also demonstrated a 1-year incidence of de-novo CKD estimated at greater than 25%. A higher proportion of African American patients developed wRF. Among those who developed wRF lower burden of prior CKD or IHD, worse NYHA class, and lower rates of SGLT-2i prescription were recorderd.

Compared to contemporary data, the development of renal dysfunction in our study is consistent with prior analyses. In a French study of 103 vATTR-CA patients, almost one-third developed CKD within a follow-up of almost 8 years (4). In another study from Italy of patients with hATTR with a median follow-up time of 56 months, kidney dysfunction was present in 15% of the patients, with a similar Portuguese study of 403 symptomatic patients with the V30M mutation estimating CKD prevalence at 36% (6,7). Recent post-hoc analysis of the ATTR-ACT study, which included both the wtATTR and hATTR, demonstrated deteriorating renal function over a 30-month period (9). In comparison, our study investigated short-term decline in renal function during the first year following diagnosis of ATTR-CA and found relatively high rates of renal dysfunction. While most available data have been derived from the hATTR population and renal dysfunction has been primarily associated with the hATTR phenotype (4–7,10), our study encompassed both wild-type and hereditary ATTR-CA phenotypes and found no significant differences in rates of developing de-novo CKD between these two groups.

Racial variability was noted, with a higher proportion of African American patients developing wRF over time in our cohort. This might be attributed to less access to care and follow-up among African American patients, resulting in later disease diagnosis (11). Published data on ATTR-CA revealed that African Americans were more likely to present with impaired systolic function at the time of diagnosis (12). In addition, African Americans are susceptible to a higher prevalence and earlier onset of chronic diseases than other racial groups (13). They also have a higher prevalence of other comorbidities, such as hypertension, which tend to present at younger ages and are 2-fold more likely to develop diabetes. Such comorbidities are well-known to significantly impact renal outcomes among patients with heart failure (14). The current study lends insight into ongoing efforts to further address disparate heart failure outcomes in minority populations, and further dedicated long-term studies should be considered to investigate renal outcomes among minorities with ATTR-CA.

Another key finding of this study is that a lack of SGLT-2i therapy is a significant predictor of wRF in this patient population. With the use of other common heart failure therapies limited by detrimental effects such as symptomatic hypotension with the use of ACEi and ARB, negative inotropic and chronotropic effects with beta-blockers, and the potential for irreversible binding to amyloid fibrils potentiating drug toxicities with calcium channel blockers and digoxin, expanding the spectrum of available systemic pharmacologic agents in improving renal outcomes is vital (15). Numerous recent studies have shown the benefit of SGLT-2i therapy in patients with HFpEF physiology as well as those with CKD through a variety of mechanisms that remain incompletely understood (15–20). To our knowledge, our study represents the first demonstration of a renal-protective effect of SGLT-2i in a mixed population of ATTR-CA patients of hATTR and wtATTR subtypes alike. Further studies are needed to validate this association and help assess whether SGLT2-i use among patients with ATTR-CA should become standard of care, particularly considering the lack of other evidence-based therapies among this difficult-to-manage population.

This study also found that wRF in our cohort was more evident among those without ischemic heart disease. This could be attributed to other ongoing systemic pathophysiologies that may contribute to a faster decline in renal function. Additionally, patients with no history of CKD were more likely to develop wRF than those with a prior history of CKD, and the direct and indirect impact of ATTR-CA on renal function in this cohort seemed to be more pronounced among these patients. Therefore, we investigated the burden of renal dysfunction among those with no prior history of CKD and estimated that a quarter of the patients had a new incidence of CKD during the study period. These patients were more likely to have a worse NYHA class at presentation and higher NT-proBNP, consistent with prior published studies in the overall population of HF patients and likely reflecting a state of chronic congestion, further promoting renal dysfunction (21,22). One study demonstrated that the risk for wRF, defined as an increase in serum creatinine ≥0.3 mg/dL, increased by 4.0 (95% CI 2.1-7.5) for each standard deviation of log NT-proBNP (21). In addition, these patients who developed CKD had lower LVEF at baseline, highlighting the burden of cardio-renal interactions on the development of renal dysfunction among those with impaired ventricular function who were diagnosed with ATTR-CA in the late course of the disease progression. Also, those who developed CKD were more likely to be on loop diuretics at the time of diagnosis. These findings were consistent with a sub-analysis of the Effect of N-3 Polyunsaturated Fatty Acids in Patients with Chronic Heart Failure (GISSI-HF) trial, demonstrating an earlier and steeper decline in eGFR among HF patients who were prescribed loop diuretics (23). This suggests that a multi-disciplinary approach is warranted when addressing patients with ATTR-CA and volume overload and requires balancing diuresis while also monitoring for hemodynamic alterations that might reduce renal perfusion (1).

Finally, the influence of concurrent tafamidis therapy on renal outcomes in our cohort was considered. While the impact of tafamidis on renal outcomes in patients with ATTR-CA has not been widely investigated, a post-hoc analysis of the ATTR-ACT revealed a slower decline and improvement in CKD staging in those who were prescribed this drug representing a potential source for confounding (9). Our population featured a high rate of tafamidis prescription, though rates of prescription were comparable across the wRF and mRF cohorts with no evident discrepancy between the two groups.

## Limitations

Our study has several limitations. This was a retrospective study with non-standardized follow-up dates, limiting standardization capability across our presented population. Further, serum creatinine was used to evaluate kidney function which may lend toward an overestimation of GFR, given general muscle wasting is expected in patients with amyloidosis. In addition, other indicators of wRF, such as proteinuria/microalbuminuria, cystatin c, and free light chain measurements, which may represent more sensitive indicators of the early stages of renal dysfunction, were not included in this study (1,3). Regardless of these limitations, the study included a relatively large overall population size with longitudinal follow-up.

## Conclusion

Our study demonstrated that the development of new or worsening renal dysfunction is common following the diagnosis of ATTR-CA. Patients with both wtATTR and hATTR were at similar risk. The wRF group had higher rates of African American race, and lower burden of prior CKD or IHD. Additionally, we identified worse NYHA class and no prior history of IHD as significant predictors associated with developing wRF, while receiving SGLT2i therapy appeared to be protective in this population (Figure 2).

**Figure 2.**
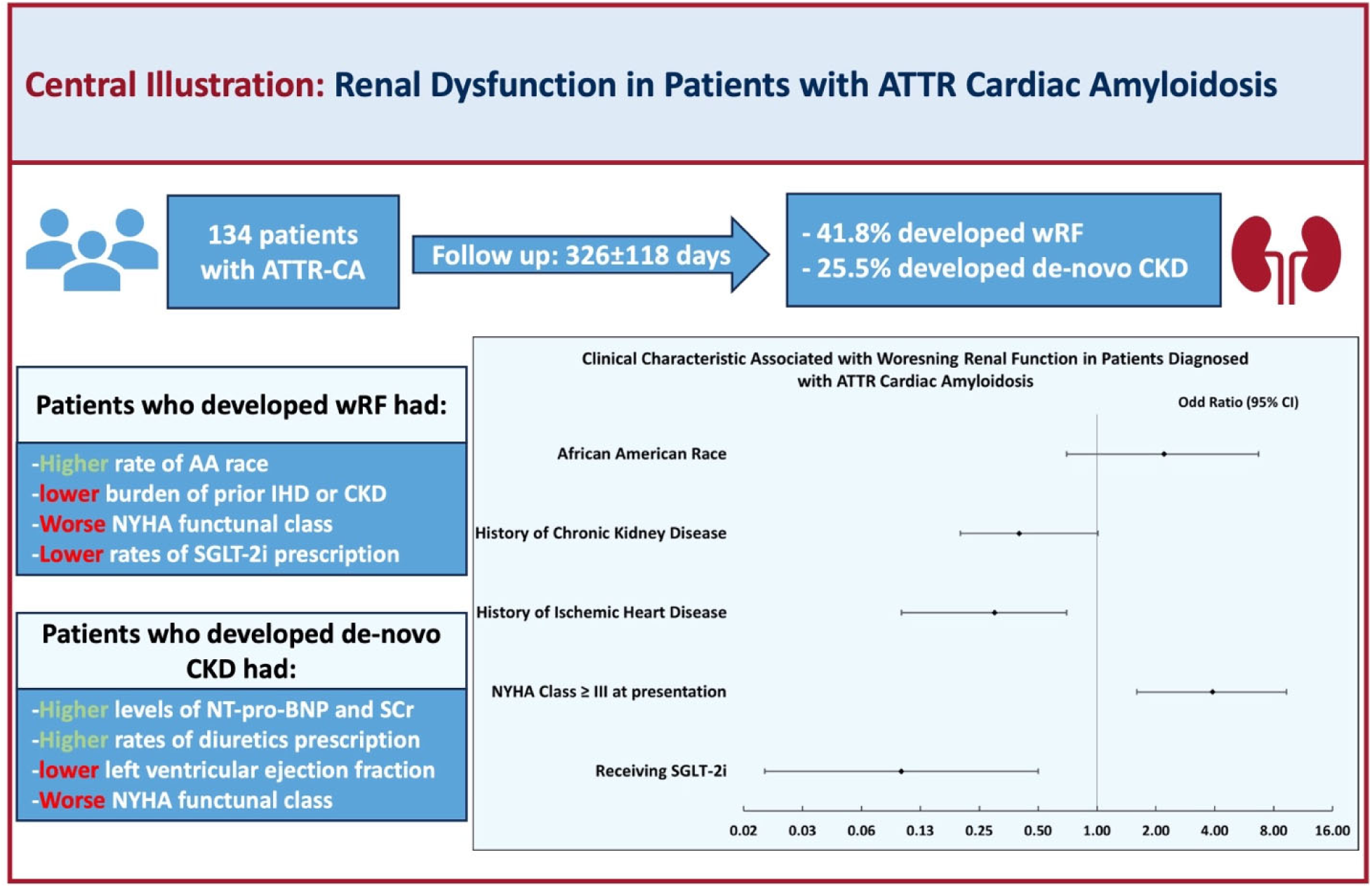
Central Illustration: Renal Dysfunction in Patients with ATTR Cardiac Amyloidosis. *AA: African American, ATTR-CA: transthyretin cardiac amyloidosis, CKD: Chronic Kidney Disease, IHD: ischemic Heart Disease, NT-proBNP: N-terminal prohormone of Brain Natriuretic Peptide, NYHA: New York Heart Association, SCr; Serum Creatinine, SGLT-2i: sodium-glucose cotransporter-2 inhibitors. wRF: worsening renal function.

## Data Availability

Data supporting this study are included within the article and/or supporting materials.

## Abbreviations

ATTR-CA: transthyretin cardiac amyloidosis
CKD: Chronic Kidney Disease
IHD: ischemic heart disease
mRF: maintained renal function.
NT-proBNP: N-terminal prohormone of Brain Natriuretic Peptide
NYHA: New York Heart Association
SGLT-2i: sodium-glucose cotransporter-2 inhibitors
wRF: worsening renal function.

## Acknowledgments

None

